# COVID-19 higher morbidity and mortality in Chinese regions with lower air quality

**DOI:** 10.1101/2020.05.28.20115832

**Authors:** Riccardo Pansini, Davide Fornacca

## Abstract

We investigated the geographical character of the COVID-19 infection in China and correlated it with satellite- and ground-based measurements of air quality. Controlling for population size, we found more viral infections in those areas afflicted by high Carbon Monoxide, formaldehyde, PM 2.5, and Nitrogen Dioxide values. Higher mortality was also correlated with relatively poor air quality. Air pollution appears to be a risk factor for the incidence of this disease, similar to smoking. This suggests the detrimental impact of air pollution in these types of respiratory epidemics.

**Short summary:**

- There is a significant correlation between air pollution and COVID-19 spread and mortality in China.
- The correlation stands at a second-order administration level, after controlling for varying population densities and removing Wuhan and Hubei from the dataset.
- Living in an area with low air quality is a risk factor for becoming infected and dying from this new form of coronavirus.

## To the Editor

COVID-19, initially detected in China and rapidly spread to the rest of the world, has ignited a pandemic at an exorbitant human cost. In a span of just over three months, eastern and western doctors, biologists, and sociologists alike have turned their attention at disentangling the aetiology of this disease. Various risk factors have been implicated with the fast spread of the virus, assuming different characters whether considered within or between countries. On an individual level, an older age, the male gender and smoking status have all been shown to increase the virulence of SARS-CoV-2.

From the standpoint of the natural sciences and evolution, we can take a broader perspective beyond virological and medical mechanisms to appreciate how a coronavirus transmitted once more from an animal species to us. Elements including human and livestock overpopulation, biodiversity loss and climate change played a critical role in making the ground suitable for a new epidemics to flourish^1^.

Pertaining to climate change, air pollution is notoriously known to cause health problems and, in particular, viral respiratory infections and pneumonia to individuals exposed for several days a year^2^.

From our perspective as biologists, economists and geographers, we have investigated the expansion of the infection in China (with Hong Kong and Taiwan included – data from the Chinese government health commission) and we have correlated it with the annual tropospheric column measurements of several air pollutants sampled from the Sentinel-5 satellite time series (data from the European Space Agency portal) as well as data derived from traditional air quality stations. The data were updated on 23 May 2020, well after the first, main infection wave and they include the major 17 April update (at the time, an increase of 1,290 casualties, about 50% from the previous figure, and an increase of 325 infections for the city of Wuhan only). See Table 1 S.I. for a comprehensive summary of the data and the sources.

Controlling for varying population sizes, we find more viral infections and fatalities in those Chinese prefectures afflicted by common pollutants of the air: CO, HCHO, PM 2.5, PM 10, and NO_2_. This trend holds also after removing (1) Wuhan city and (2) the whole Hubei province from the dataset in succession (see Table 4 S.I. and 5 S.I.). All correlation tests were performed using Kendall’s Tau with the significance threshold set to 0.05.

Aerosol data from the satellite, which include PM 2.5 and PM 10, were not associated with higher mortality. This is not surprising, they in fact comprise non-pollutant, inert particulates such as dust, sand and sea salt. On the other hand, statistics suggested that higher levels of O_3_ and SO_2_ were not associated with more COVID-19 deaths, which goes against the trends of the other pollutants, an aspect that requires further investigation. A comprehensive statistical output is reported in Table 1.

**Table 1.** Correlation between satellite- and ground-based air quality variables and cumulated COVID-19 infections per 100,000 inhabitants, fatalities per 100,000 inhabitants, and mortality rate in China, until 23 May 2020.

| China | Infections<br>(/100k pop) |  |  | Fatalities<br>(/100k pop) |  |  | Mortality<br>(fatalities/infections) |  |  |
| --- | --- | --- | --- | --- | --- | --- | --- | --- | --- |
|  | df (N-2) | tau | p-value | df (N-2) | tau | p-value | df (N-2) | tau | p-value |
| CO sat | 337 | .28 | <.001 | 337 | .19 | <.001 | 313 | .16 | <.001 |
| NO<sub>2</sub> sat | 337 | .23 | <.001 | 337 | .14 | .001 | 313 | .12 | .006 |
| O<sub>3</sub> sat | 337 | -.08 | .030 | 337 | .00 | .967 | 313 | .02 | .635 |
| SO<sub>2</sub> sat | 337 | -.10 | .005 | 337 | -.02 | .634 | 313 | .00 | .964 |
| Aerosol sat | 337 | -.12 | .001 | 337 | -.03 | .488 | 313 | .00 | .950 |
| HCHO sat | 337 | .34 | <.001 | 337 | .20 | <.001 | 313 | .17 | <.001 |
| PM 2.5 ground | 302 | .15 | <.001 | 302 | .18 | <.001 | 285 | .18 | <.001 |
| PM 10 ground | 302 | .04 | .330 | 302 | .12 | .006 | 285 | .13 | .005 |
| CO ground | 302 | -.01 | .840 | 302 | .11 | .012 | 285 | .12 | .007 |
| NO<sub>2</sub> ground | 302 | .12 | .002 | 302 | .12 | .005 | 285 | .12 | .007 |
| O<sub>3</sub> ground | 302 | -.03 | .477 | 302 | -.02 | .585 | 285 | -.03 | .482 |
| SO<sub>2</sub> ground | 302 | -.01 | .843 | 302 | .04 | .409 | 285 | .06 | .178 |
| population | 337 | .23 | <.001 | 337 | .17 | <.001 |  |  |  |
| pop density | 337 | .32 | <.001 | 337 | .16 | <.001 |  |  |  |

Despite the fact the SARS-CoV-2 was first detected in Wuhan and that the first location of the pathogen assumes a key role in the geographical spread of the infection, the detrimental effect that air pollution assumes at a national level remains evident. Although the measures to contain the virus taken by the Chinese government were effective when compared to other countries affected by what later became a pandemic, the strong association between air pollution and higher morbidity and mortality rates can be detected even where the series of outbreaks originated from.

Even if free health care was dispensed to everyone in the exceptional case of this epidemics, the Chinese health system, as those ones of many other countries, is not adequate at planning for global health according to these risk factors^3^. We hypothesise that the correlational significance we have found points towards air pollution as a critical risk cofactor for COVID-19, also because of other studies. (1) Testing the more proximal hypothesis that COVID-19 outbreaks could follow with a temporal delay from days with high NO_2_ presence in the air, colleagues in Shanghai are publishing detailed time series suggesting a delay of 12 days before hospitalisations for the Hubei province^4^. This may even suggest the role of air pollutants as airborne vectors for this virus, later also suggested by another study^5^ run in three cities in Hubei province. At a Chinese national level, PM 2.5 and NO_2_ pollution from ground stations was recently found correlated with this pathology incidence, after adjusting for some socio-economic factors and human movements following the Spring festival^6^. (2) In the United States, chronic exposure to high levels of PM 2.5 have been found co-responsible for a higher mortality rate from respiratory diseases, specifically in 2020, that is in the presence of COVID-19^7^. This is a rate higher than 11 other American demographic co-variables. (3) In Italy, a similar positive correlation controlling for five demographic variables was also reported^8^, with the additional and novel evidence that fragments of the RNA from this virus were found in the particulate matter of the harshly hit northern Italian city of Bergamo^9^, laying in the most polluted European area of the Po valley. (4) Finally, our comprehensive study including eight countries assessed with a similar analysis, confirms this same trend in seven additional countries to China (Italy, United States, Iran, Spain, France, Germany, U.K.).

As a clear and immediate action to prevent the trajectory of this and future epidemics, curbing climate change^10^ must be endorsed way more seriously. With no exceptions, it must be endorsed now. Will the smallest of the parasites be able to wake us up this time, so that we start caring about the health of the environment as much as we have been caring about our own and public health?

## Data Availability

The data are available in github.com

## Acknowledgements

No funding was obtained for the completion of this study. Livia Ottisova improved and revised the manuscript. Chun Chen commented on statistics. RP did this work while in isolation in the Italian Po valley, due to the ongoing pandemic.

---

**Figure 1.**
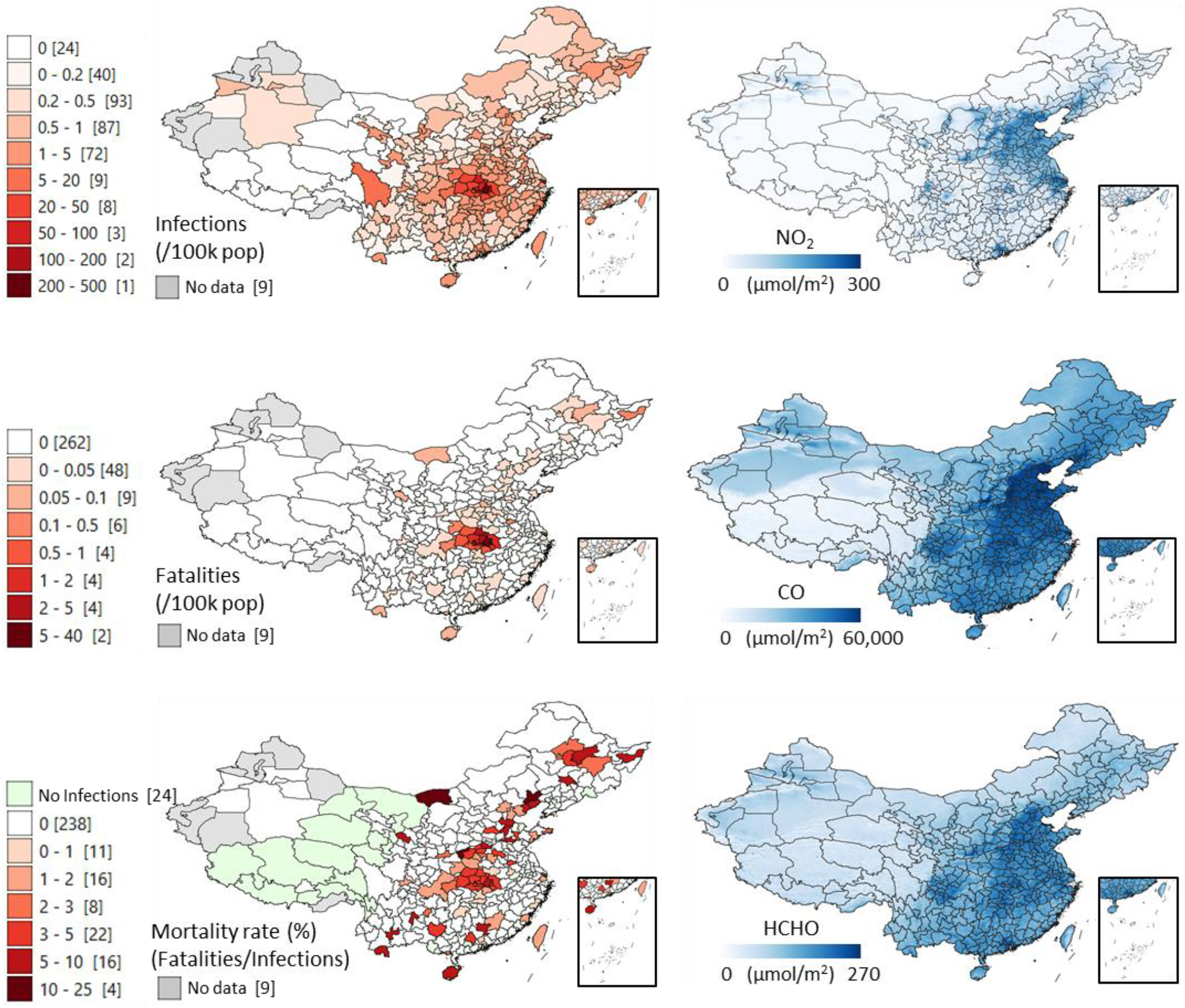
Distribution of COVID-19 infections, fatalities and mortality rates across the prefectures of China (updated on 23 May 2020), and the distribution of the tropospheric column amounts of three representative air pollutants derived from satellite (2019 averages): Nitrogen Dioxide (NO_2_), Carbon Monoxide (CO), and Formaldehyde (HCHO). The values in the square brackets show the COVID-19 cases’ counts of administrative units.

## Digital supplement

**Table 1 S.I.** The datasets information with their sources.

| Data | Measuring Unit | Time period | Format | Source |
| --- | --- | --- | --- | --- |
| COVID-19 | No. of infections,<br>No. of deaths | Updated on<br>23 May 2020 | Tabular<br>Prefecture level | DXY - DX Doctor:<br>http://ncov.dxy.cn/ncovh5/view/en_pneumonia<br>from Chinese government health commission |
| Population | No. of residents | Estimates<br>2017 | Tabular<br>Prefecture level | https://www.citypopulation.de/<br>Data from provincial governments |
| Air quality, ground stations: |  |  |  |  |
| PM2.5, PM10, O<sub>3</sub>,<br>NO<sub>2</sub>, SO<sub>2</sub>, CO | AQI | 2014 | Tabular<br>GPS points | University of Harvard Dataverse:<br>https://dataverse.harvard.edu<br>Data from http://aqicn.org |
| Air quality, satellite: |  |  |  |  |
| UV Aerosol Index<br>CO, HCHO, NO<sub>2</sub>, O<sub>3</sub>,<br>SO<sub>2</sub> | Qualitative Index<br>mol/m<sup>2</sup> | 2019 | Continuous grid<br>(0.01 arc deg.) | Sentinel-5 Atmospheric variables<br>https://developers.google.com/earth-engine/datasets/tags/air-quality |

**Table 2 S.I.**
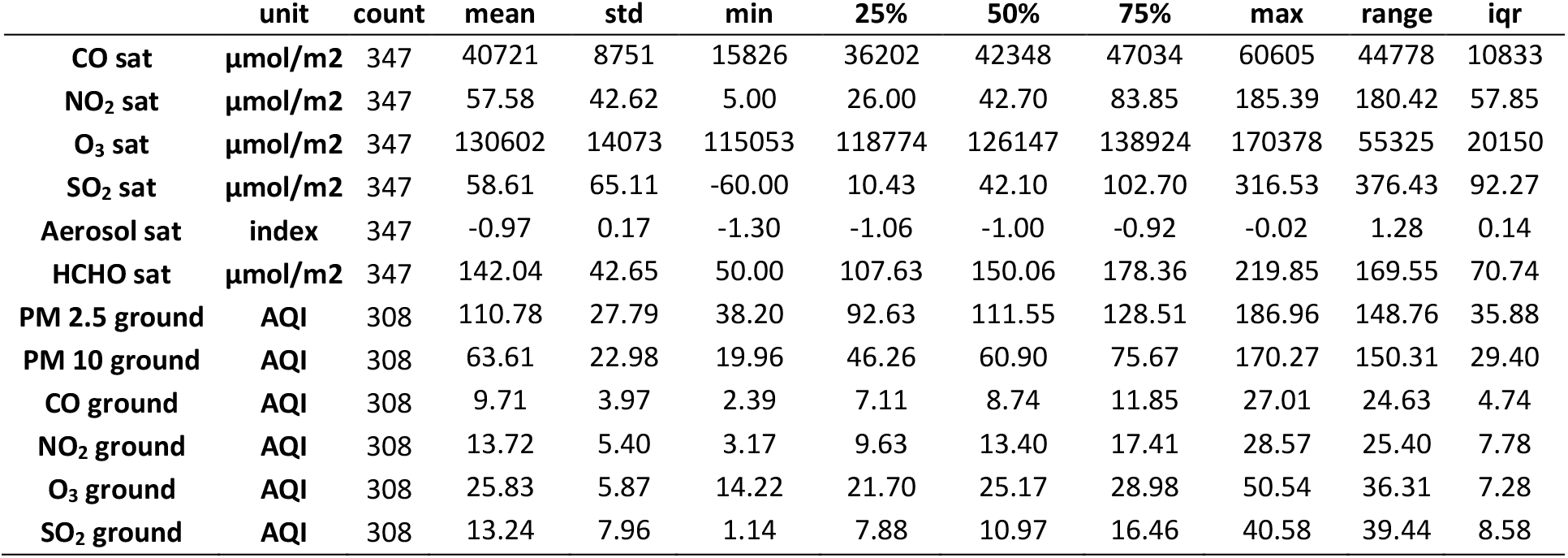
Descriptive statistics for the different satellite- and ground-based air quality measurements.

**Table 3 S.I.** Correlation between satellite- and ground-based air quality variables and population density in China.

|  | Population density |  |  |
| --- | --- | --- | --- |
|  | df (N-2) | tau | p-value |
| CO sat | 345 | .39 | <.001 |
| NO<sub>2</sub> sat | 345 | .37 | <.001 |
| O<sub>3</sub> sat | 345 | .36 | <.001 |
| SO<sub>2</sub> sat | 345 | .36 | <.001 |
| Aerosol sat | 345 | -.07 | .039 |
| HCHO sat | 345 | .00 | .890 |
| PM 2.5 ground | 345 | -.07 | .046 |
| PM 10 ground | 345 | .40 | <.001 |
| CO ground | 306 | .24 | <.001 |
| NO<sub>2</sub> ground | 306 | .13 | .001 |
| O<sub>3</sub> ground | 306 | .04 | .327 |
| SO<sub>2</sub> ground | 306 | .30 | <.001 |

**Table 4 S.I.** Correlation between satellite- and ground-based air quality variables and cumulated COVID-19 infections per 100,000 inhabitants, fatalities per 100,000 inhabitants, and mortality rate in China (without the administrative unit of Wuhan), until 23 May 2020.

| <u>China without</u><br><u>Wuhan</u> | <u>Infections</u><br>(/100k pop) |  |  | <u>Fatalities</u><br>(/100k pop) |  |  | <u>Mortality</u><br>(fatalities/infections) |  |  |
| --- | --- | --- | --- | --- | --- | --- | --- | --- | --- |
|  | df (N-2) | tau | p-value | df (N-2) | tau | p-value | df (N-2) | tau | p-value |
| CO sat | 336 | .28 | <.001 | 336 | .18 | <.001 | 312 | .16 | <.001 |
| NO<sub>2</sub> sat | 336 | .22 | <.001 | 336 | .14 | .001 | 312 | .12 | .008 |
| O<sub>3</sub> sat | 336 | -.08 | .031 | 336 | .00 | .943 | 312 | .02 | .617 |
| SO<sub>2</sub> sat | 336 | -.10 | .006 | 336 | -.02 | .690 | 312 | .00 | .975 |
| Aerosol sat | 336 | -.12 | .001 | 336 | -.03 | .537 | 312 | .00 | .992 |
| HCHO sat | 336 | .34 | <.001 | 336 | .19 | <.001 | 312 | .16 | <.001 |
| PM 2.5 ground | 301 | .15 | <.001 | 301 | .18 | <.001 | 284 | .17 | <.001 |
| PM 10 ground | 301 | .04 | .361 | 301 | .12 | .008 | 284 | .13 | .006 |
| CO ground | 301 | -.01 | .798 | 301 | .11 | .013 | 284 | .12 | .008 |
| NO<sub>2</sub> ground | 301 | .11 | .003 | 301 | .12 | .009 | 284 | .11 | .012 |
| O<sub>3</sub> ground | 301 | -.03 | .512 | 301 | -.02 | .637 | 284 | -.03 | .528 |
| SO<sub>2</sub> ground | 301 | -.01 | .851 | 301 | .04 | .396 | 284 | .06 | .171 |
| population | 336 | .22 | <.001 | 336 | .16 | <.001 |  |  |  |
| pop density | 336 | .32 | <.001 | 336 | .15 | <.001 |  |  |  |

**Table 5 S.I.** Correlation between satellite- and ground-based air quality variables and cumulated COVID-19 infections per 100,000 inhabitants, fatalities per 100,000 inhabitants, and mortality rate in China (without Hubei province), until 23 May 2020.

| <u>China without</u><br><u>Hubei prov.</u> | <u>Infections</u><br>(/100k pop) |  |  | <u>Fatalities</u><br>(/100k pop) |  |  | <u>Mortality</u><br>(fatalities/infections) |  |  |
| --- | --- | --- | --- | --- | --- | --- | --- | --- | --- |
|  | df (N-2) | tau | p-value | df (N-2) | tau | p-value | df (N-2) | tau | p-value |
| CO sat | 320 | .26 | <.001 | 320 | .13 | .003 | 296 | .11 | .013 |
| NO<sub>2</sub> sat | 320 | .26 | <.001 | 320 | .17 | <.001 | 296 | .15 | .001 |
| O<sub>3</sub> sat | 320 | .26 | <.001 | 320 | .13 | .002 | 296 | .11 | .011 |
| SO<sub>2</sub> sat | 320 | .24 | <.001 | 320 | .15 | .001 | 296 | .13 | .005 |
| Aerosol sat | 320 | -.07 | .054 | 320 | .03 | .551 | 296 | .04 | .417 |
| HCHO sat | 320 | -.08 | .044 | 320 | .04 | .386 | 296 | .05 | .275 |
| PM 2.5 ground | 320 | -.10 | .008 | 320 | .03 | .539 | 296 | .05 | .299 |
| PM 10 ground | 320 | .32 | <.001 | 320 | .14 | .002 | 296 | .11 | .016 |
| CO ground | 286 | .10 | .011 | 286 | .11 | .018 | 269 | .11 | .022 |
| NO<sub>2</sub> ground | 286 | .00 | .992 | 286 | .07 | .108 | 269 | .08 | .087 |
| O<sub>3</sub> ground | 286 | -.03 | .395 | 286 | .09 | .052 | 269 | .10 | .032 |
| SO<sub>2</sub> ground | 286 | .13 | .001 | 286 | .15 | .002 | 269 | .14 | .003 |
| population | 320 | .26 | <.001 | 320 | .21 | <.001 |  |  |  |
| pop density | 320 | .35 | <.001 | 320 | .19 | <.001 |  |  |  |

